# Relationship between branched chain amino acids and type 2 diabetes: a bidirectional Mendelian Randomization study

**DOI:** 10.1101/2023.05.23.23290391

**Authors:** Jonathan D. Mosley, Mingjian Shi, David Agamasu, Nataraja Sarma Vaitinadin, Venkatesh Murthy, Ravi V. Shah, Minoo Bagheri, Jane F. Ferguson

## Abstract

**Background:** Human genetic studies suggest that the branched chain amino acids (BCAA) valine, leucine and isoleucine have a causal association with type 2 diabetes. However, inferences are based on analyses of a limited number of genetic loci associated with BCAAs. Whether these conclusions are supported when using instrumental variables for BCAAs that capture a broad set of genetic mechanisms is not known.

**Methods:** We constructed and validated instrumental variables for each BCAA using large well-powered datasets and tested their association with type 2 diabetes using the two-sample inverse variance weighted (IWV) Mendelian randomization (MR) approach. Sensitivity analyses were performed to ensure the accuracy of the findings. Instrumental variables for type 2 diabetes, fasting insulin and body mass index (BMI) were also tested for associations with BCAA levels.

**Results:** There were no significant associations with diabetes for valine (beta=0.17 change in log-odds per standard deviation change in valine, [95% CI, −0.28 - 0.62], p=0.45), leucine (beta=0.19 [−0.30 - 0.68] p=0.45) or isoleucine (beta=0.02 [−0.54 - 0.59], p=0.94). In contrast, type 2 diabetes was associated with each BCAA (valine: beta=0.08 per standard deviation change in levels per log-odds change in type 2 diabetes, [0.05 - 0.10], p=1.8×10^−9^), (leucine: beta= 0.06 [0.04 - 0.09], p=4.5×10^−8^) and isoleucine (beta= 0.06 [0.04 - 0.08], p=2.8×10^−8^). The type 2 diabetes associations were replicated in an independent population, but not in a second population where type 2 diabetes cases were removed, highlighting the consistency and specificity of the association. Similar positive associations were seen for fasting insulin and BMI with the BCAAs. In multivariable MR analyses, type 2 diabetes and fasting insulin had consistent independent associations with each BCAA.

**Conclusions:** These data suggest that the BCAAs are not mediators of type 2 diabetes risk but are biomarkers of diabetes and higher insulin.

## INTRODUCTION

The branched chain amino acids (BCAA) valine, leucine and isoleucine are essential amino acids with functional roles that include regulation of lipid, glucose and protein metabolism.^1^ It has long been recognized that BCAA levels in blood are elevated in individuals with impaired insulin secretion and obesity.^2^ These early observations have prompted interest as to whether the BCAAs may have an etiological role in insulin resistance and metabolic disease, including type 2 diabetes. Human studies suggesting roles for the BCAAs as upstream drivers of metabolic disease include a prospective study of normoglycemic participants that found individuals with the highest baseline BCAA levels were at 5-fold increased risk of incident diabetes.^3^ However, findings from epidemiological studies have not been consistent in all populations.^4–8^

Support for an etiological role of the BCAAs in the development of type 2 diabetes has also come from human genetics studies. Genetic methodologies, such as Mendelian randomization, use single nucleotide polymorphisms (SNPs) as instrumental variables to determine whether an exposure may have a causal association with an outcome.^9^ An advantage of these studies is that they can be less prone to biases such as reverse causation, which can lead to incorrect inferences in epidemiological studies.^10^ One study focused largely on SNPs flanking the gene *PPM1K*, which encodes a protein phosphatase which activates the branched chain α-ketoacid dehydrogenase complex (BCKDC) that catalyzes the first irreversible step in the BCAA metabolism.^11^ The investigators identified a significant positive association between genetically predicted BCAA levels and type 2 diabetes risk, suggesting that BCAAs are upstream mediators of risk.^12^ A subsequent study examined 3 genetic instruments near the *PPM1K, SLC1A4, ASGR1* genes and did not find an association with Homeostatic Model Assessment for Insulin Resistance (HOMA-IR).^13^ Rather they found a polygenic predictor of HOMA-IR was positively associated with the BCAAs, suggesting that the BCAAs are elevated as a consequence of insulin resistance. These results were supported by a later study showing a positive association between genetic predictors of insulin resistance phenotypes and BCAAs. A common unifying interpretation of these findings is that BCAAs may be the causal down-stream mediators linking insulin resistance and obesity phenotypes to type 2 diabetes risk.^12,14^

A limitation of the prior genetic studies is that they examined a small number of genetic loci associated with BCAA levels, thereby limiting the strength of evidence for a causal role of the BCAAs on type 2 diabetes. Recent large-scale genome-wide association studies (GWAS) of BCAAs have identified considerably more genetic loci associated with the BCAAs, providing an opportunity to investigate etiological relationships using robust predictors that represent heterogenous genetic mechanisms modulating BCAA levels.^15^ We developed genetic instruments for the BCAAs and metabolic phenotypes using data derived from contemporary large-scale GWAS and applied Mendelian randomization approaches to define the relationships among these phenotypes.

## METHODS

### GWAS Summary Statistics data sets

Single nucleotide polymorphisms (SNPs) associated with valine, leucine and isoleucine used in the primary analyses were derived from GWAS summary statistics generated by a study examining 115,082 UK Biobank European Ancestry participants with NMR metabolomics from Nightingale Health (biomarker quantification version 2020).^15^ Prior to GWAS, metabolite levels were inverse rank-normalized and set to a standard deviation of 1. Data were downloaded from the NHGRI-EBI GWAS Catalog (GCST90092891, GCST90092843, GCST90092995).^16^ GWAS summary statistics used for validation of SNP genetic instruments were obtained from the Metabolic Syndrome in Men (METSIM) consortium, a study of 6136 Finnish men without prevalent or incident type 2 diabetes and with metabolites measured using the relative quantitative liquid chromatography–tandem mass spectrometry (LC-MS/MS) Metabolon DiscoveryHD4 platform.^17^ Metabolite level residuals were inverse-normalized and summary statistics were downloaded from https://pheweb.org/metsim-metab/. Summary statistics were also obtained a study of 7,824 European ancestry participants from the TwinsUK and Cooperative Health Research in the Region of Augsburg (KORA) cohort who underwent metabolite acquisition by Metabolon using GC-MS or UPLC-MS/MS. Summary statistics based on log10 transformed metabolite levels were obtained from http://metabolomics.helmholtz-muenchen.de/gwas/.18

Summary statistics for type 2 diabetes from a study of 898,130 cases and controls of European ancestry were obtained from the DIAGRAM consortium (https://diagram-consortium.org/downloads.html.19 Summary statistics were from a subset of the cohort that did not include the UK Biobank population. Summary statistics of log-transformed fasting insulin from a study of 151,188 individuals of European ancestry without type 2 diabetes^20^ were obtained from the MAGIC consortium (https://magicinvestigators.org/downloads/). Summary statistics for body mass index (BMI) were from a study of 339,224 adults of European ancestry and obtained from the GIANT consortium (https://portals.broadinstitute.org/collaboration/giant/index.php/GIANT_consortium_data_files).^2^ ^1^ None of these data sets included participants from the UK Biobank to ensure there was no overlap with the UK Biobank metabolite cohort.

### Analysis

#### Lead SNP identification

Lead SNPs associated with valine, leucine and isoleucine were selected from the UK Biobank summary statistics using a clumping algorithm that selected a linkage disequilibrium (LD)-reduced set of SNPs (r-squared <0.001, selection window of 3000 Kb) and a minor allele frequency (MAF) >1%.

#### Annotation

The FUMA webserver (https://fuma.ctglab.nl/), with default settings, was used to identify nearby genes and expression QTLs (based on GTEx v8.0) associated with candidate SNPs.^22^ The DAVID program was used to identify ontologies, cellular locations and pathways associated with candidate genes.^23,24^ These data in conjunction with a manual literature review was used to identify the likely candidate genes located near associated SNPs. If a single likely candidate gene could not be identified, all plausible candidate genes are presented. Visualization of the cellular locations of candidate genes were created with BioRender.com.

#### Mendelian Randomization

Mendelian randomization (MR) approaches were used to test associations between genetic instruments for the BCAAs and outcomes. MR is an analytic framework that uses SNPs associated with diverse pleiotropic mechanisms as instrumental variables to test the association between an exposure and outcome.^9,10^ The validity of the approach relies on the following assumptions: (1) Relevance: the SNP instrumental variables are associated with the exposures; (2): Independence: the instrumental variables are not associated with confounders; and (3) Exclusion restriction: the instrumental variables association with the outcome is only through the exposure.^25^ We demonstrate the relevance assumption by validating that our BCAA genetic instruments associated with their corresponding BCAAs in an independent population. We conducted a range of MR sensitivity analyses, as well as multivariable analyses to decrease the likelihood of violating the other assumptions.

SNP genetic instruments were constructed for each BCAA from the UK Biobank GWAS. Non-palindromic common (MAF>5%) SNP instruments were selected using a clumping algorithm and SNPs that were available on each set of summary statistics for the METSIM BCAAs, type 2 diabetes and fasting insulin levels. Two-sample MR approach was used to test the association between BCAA and outcome (the corresponding BCAA in METSIM and type 2 diabetes) using inverse-variance weighted regression (IVW).^26^ The Cochran’s Q statistic and its associated p-value, was used to assess for horizontal pleiotropy, and p<0.05 was deemed suggestive of pleiotropy. In addition, sensitivity analyses were performed using MR-Egger to assess for horizontal pleiotropy and the weighted median method.^27,28^ A leave-one-out analyses was also performed for the BCAA/type 2 diabetes associations. Associations were tested using the Mendelian Randomization R package.^29^ MR-PRESSO was used to assess for horizontal pleiotropy (global test) and distortions in IVW estimates (distortion test) with exclusion of SNPs identified as outliers at p<0.05/(# of genetic instruments).^30^

The same MR approaches were used to assess whether instrumental variables for type 2 diabetes, fasting insulin and BMI were associated with each BCAA. The association between type 2 diabetes and each BCAA was also examined using metabolites measured in the KORA and METSIM studies.

Pairwise multivariable MR (MVMR) was used to test for whether the associations between type 2 diabetes and the BCAAs was independent of fasting insulin and BMI.

All statistical tests were two-sided and a nominal p<0.05 was considered significant.^31^

## RESULTS

We identified independent common SNPs (MAF>1%) significantly associated (p<5×10^−8^) with the BCAAs from GWAS performed on 115,082 European Ancestry UK Biobank participants. There were 18, 15 and 8 associated SNPs for valine, leucine and isoleucine, respectively (**Table 1** and **Supplemental Table 1**). Five SNPs were located near genes examined in prior BCAA association studies (*GCKR, PPM1K, DDX19A, SLC1A4* and *ASGR1* [which is near *SLC2A4*]).^12,13^ Two associations were near gene loci that have direct roles in BCAA metabolism (*PPM1K*, *BCAT2*), while others had known functionality related to intermediate metabolism (SLC1A4*, PRODH, GLS2, GLUD1, PCCB*), insulin signaling (*IRS1, GRB14, GCKR*) and glucose regulation (*GCKR, SLC2A4*) (**Figure 1**). *HNF4G* and *RLN3* have previously been associated with adiposity traits. We did not observe associations previously reported near *TRMT61A* and *CBLN1* in the UK biobank or METSIM cohorts (**Supplemental Table 2**).^12^ We constructed genetic instruments for each BCAA in the UK Biobank set (**Supplemental Table 3**). To validate these predictors, we tested their associations with corresponding BCAAs measured in the METSIM data set. Effect sizes for each SNP instrument were generally linearly related for each BCAA between the studies (**Figure 2**), and MR analyses demonstrated significant (p<0.05), positive associations for each BCAA, without heterogeneity (Cochran’s Q statistic p>0.05) (**Figure 2**, **Supplemental Table 4**).

**Figure 1:**
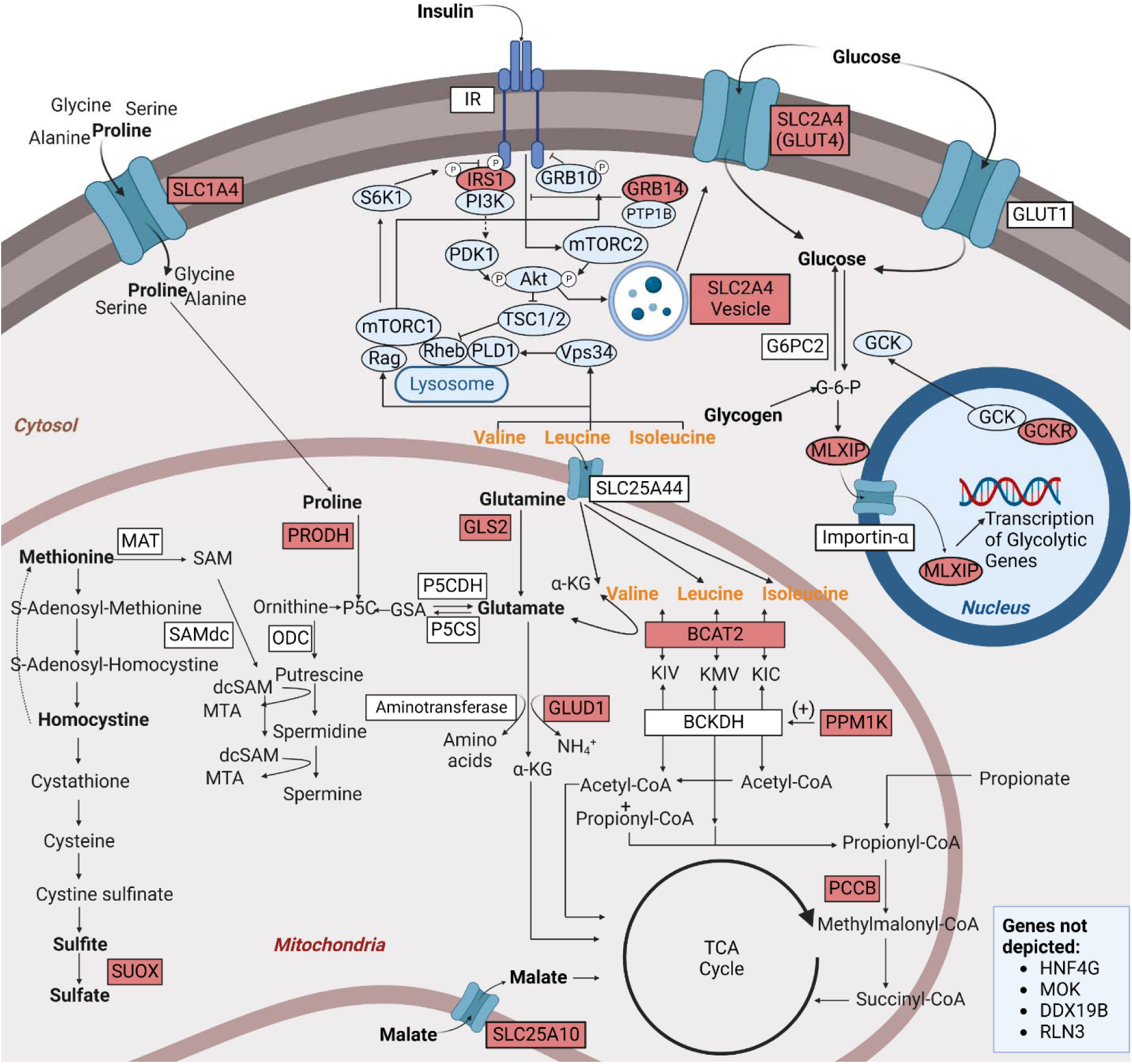
Cellular functions of proteins for genes associated with circulating valine, leucine and isoleucine levels. Genes located near lead SNPs are highlighted in red.

**Figure 2:**
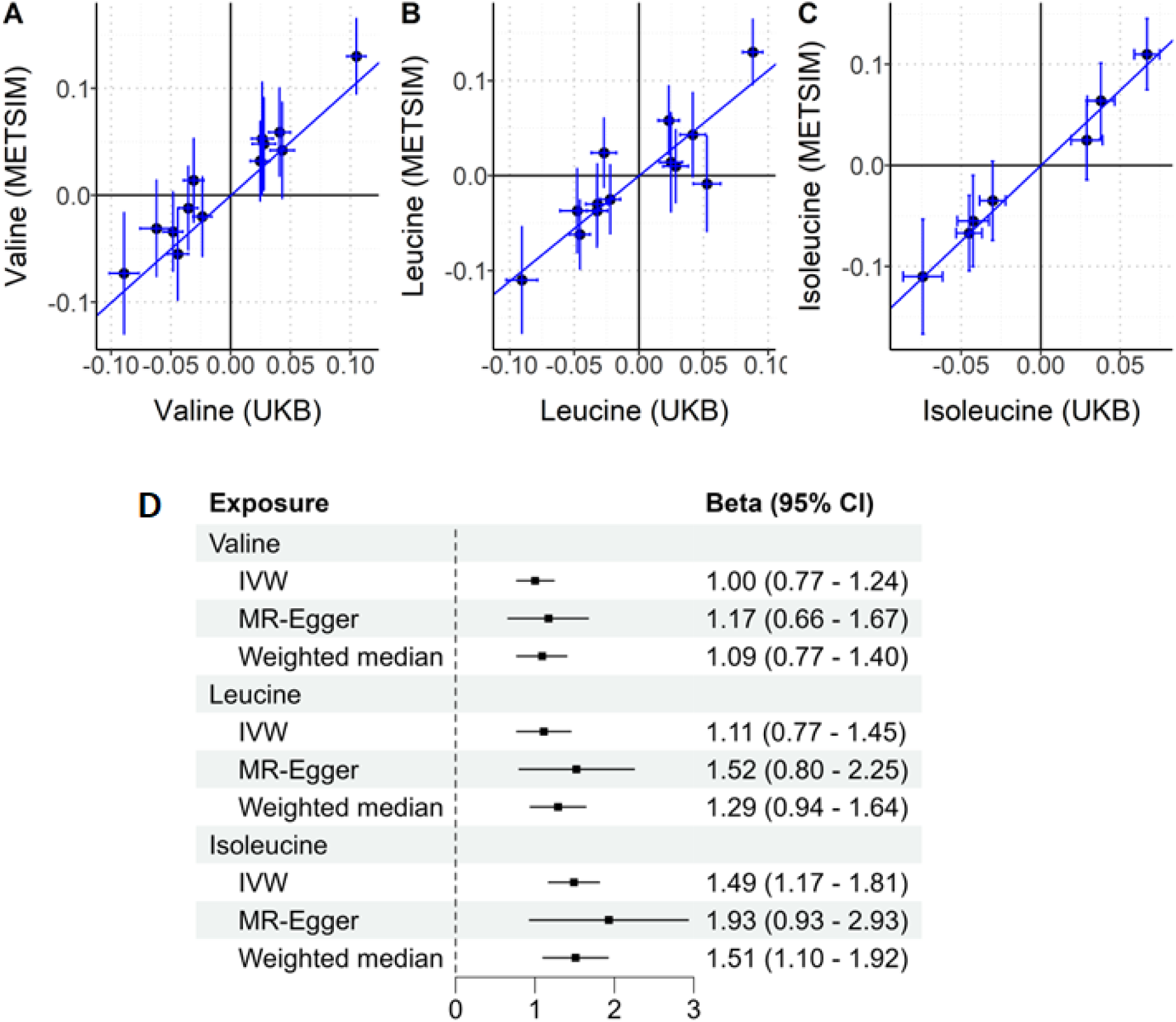
Associations between SNP instruments for the BCAAs and validation phenotypes. (A-C) Scatter plots showing comparing the GWAS effect sizes between the valine (n=13 SNPs), leucine (n=13 SNPs) and isoleucine SNP (n=7 SNPs) genetic instruments identified in the UK Biobank cohort and their corresponding amino acid in the METSIM cohort. The lines represent the association based on the IVW method. (D) Summary of associations from MR analyses testing the association between the genetic instruments in UK Biobank and their corresponding amino acid level in METSIM. IVW=inverse variance weighted method.

**Table 1.** Genomic loci and candidate genes associated with circulating valine, leucine or isoleucine levels.

| Chromosome:<br>Position<br>(Hg37) | Lead SNP(s) | Lead<br>candidate<br>gene(s)<sup>1</sup> | Function<sup>2</sup> |
| --- | --- | --- | --- |
| 2:27730940 | Val: rs1260326<br>Leu: rs1260326<br>Ile: rs1260326 | *GCKR* | Inhibits glucokinase in liver and pancreatic islet cells. |
| 2:65219030 | Val: rs2007061 | *SLC1A4* | Sodium-dependent neutral amino acid transporter for alanine, serine, cysteine, and threonine. |
| 2:165501927 -<br>2:165528876 | Val: rs13389219<br>Leu: rs5835988<br>Ile: rs5835988 | *GRB14* | Interacts with insulin receptors and inhibits insulin receptor signaling. |
| 2:227108446 | Val: rs2943652<br>Leu: rs2943652 | *IRS1* | Transmits signal between the insulin receptor and phosphoinositide 3-kinase. |
| 3:136328270 | Val: rs1471740<br>Leu: rs1471740 | *PCCB* | Breaks down amino acids including isoleucine and valine. |
| 4:88920985 | Val: rs10018448<br>Leu:<br>rs114796149,<br>rs10018448<br>Ile: rs114796149 | *PPM1K* | Activates branched chain α-ketoacid dehydrogenase complex that catalyzes the first step in the BCAA metabolism. |
| 8:76443463 -<br>8:76454025 | Leu: rs2977929<br>Ile: rs2941456 | *HNF4G* | Enables DNA-binding transcription activator activity. Associated with BMI by GWAS. |
| 10:88820592 | Val: rs17096421 | *GLUD1* | Mitochondrial matrix enzyme that catalyzes the oxidative deamination of glutamate to alpha-ketoglutarate and ammonia. |
| 12:56393337 | Val: rs1081975<br>Leu: rs1081975 | *SUOX* | Breaks down sulfur-containing amino acids. |
| 12:56861458 -<br>12:56865056 | Val: rs2638315<br>Leu: rs2638315<br>Ile: rs7302925 | *GLS2* | Catalyzes the hydrolysis of glutamine to glutamate and ammonia. |
| 12:122607112 | Val: rs12368387 | *MLXIP*<br>(*MONDOA*) | Transcriptional activation of glycolytic target genes in response to cellular glucose levels. |
| 14:102718052 | Val: rs2274815 | *WDR20*,<br>*MOK*,<br>*ZNF839* | Multiple. Lead SNP intronic in MOK. |
| 16:70270907 -<br>16:70368909 | Val: rs12325419<br>Leu: rs35452938<br>Ile: rs12325419 | *Multiple,*<br>*including*<br>*DDX19A-*<br>*DT and*<br>*DDX19B* | Multiple. Intronic in DDX19A-DT and 3' UTR in DDX19B. DDX19A identified on prior isoleucine GWAS. |
| 16:72149923 | Leu: rs9930957 | *DHODH, TX*<br>*NL4B, HP, H*<br>*PR, DHX38,*<br>*PMFBP1* | Multiple. |
| 17:7185779 | Val:<br>rs117643180<br>Leu:<br>rs117643180<br>Ile: rs117643180 | *SLC2A4*<br>*(GLUT4)* | Insulin-regulated facilitative membrane glucose transport into cells. |
| 17:79636653 | Val: rs62080209 | *SLC25A10*<br>*(DIC)* | Translocates small metabolites (e.g. malate, succinate) across the mitochondrial membrane for metabolic processes including the TCA cycle and fatty acid synthesis. |
| 19:14139004 | Leu: rs1982632 | *RLN3* | Insulin-superfamily hormone regulating appetite, food intake, and weight gain. |
| 19:49300431 -<br>19:49317459 | Val: rs35230038,<br>rs117048185<br>Leu: rs4801776<br>Ile: rs71179052 | *BCAT2* | Mitochondrial branched chain aminotransferase that catalyzes production of the BCAAs. |
| 22:18915282 -<br>22:18915347 | Val: rs2238732<br>Leu: rs5747934 | *PRODH* | Mitochondrial protein that catalyzes the first step in proline degradation. |

1. Lead candidate genes shown were selected based on their proximity to lead the lead SNP, eQTL analyses and known relevance of cellular functions to BCAA metabolism. 2. Known cellular functions for loci where a single lead candidate gene is identified.

There was no genetic association between the BCAA predictors and type 2 diabetes by IVW for valine (beta=0.17 change in log-odds per standard deviation change in valine, [95% CI, −0.28 - 0.62], p=0.45), leucine (beta=0.19 [−0.30 - 0.68] p=0.45) or isoleucine (beta=0.02 [−0.54 - 0.59], p=0.94) by the IVW method (**Figure 3**). Similar results were seen in sensitivity analyses using the leave-one-out, MR-Egger and weighted median methods (**Supplementary Figure 1**, **Supplemental Table 5**). There was significant heterogeneity (Cochran’s Q statistic p<0.05) for each association, but the MR-Egger intercept values were not significant (p>0.05), and outlier removal by the MR-PRESSO method did not identify significant distortion (p>0.05) in the effect estimates (**Supplemental Table 5**).

**Figure 3:**
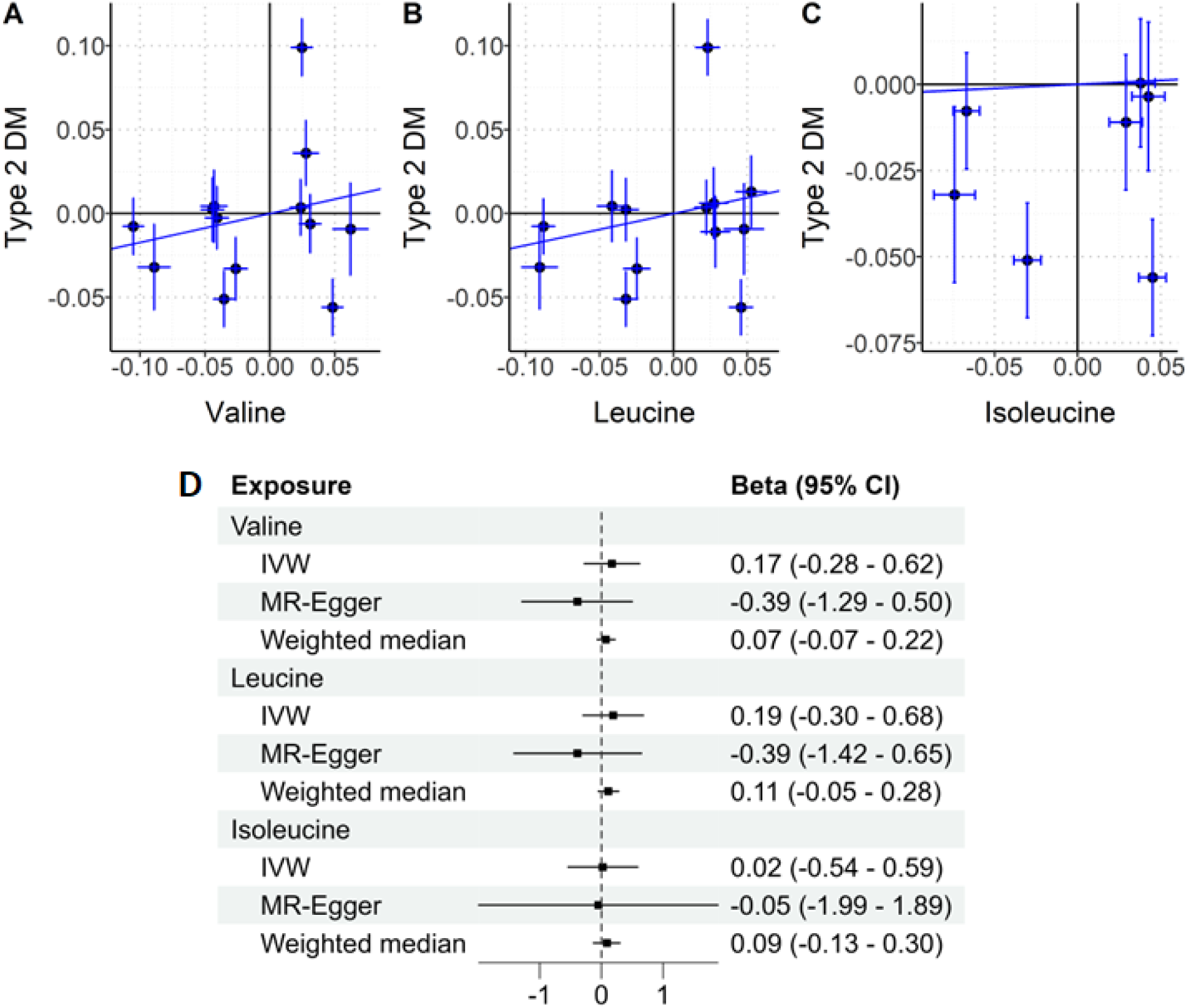
Associations between SNP instruments for the BCAAs and type 2 diabetes. (A-C) Scatter plots showing comparing the associations between the valine, leucine and isoleucine SNP genetic instruments and with type 2 diabetes. The lines represent the association based on the IVW method. (D) Summary of associations from MR analyses testing the association between the genetic instruments for each BCAA and type 2 diabetes. IVW=inverse variance weighted method.

**Figure 4:**
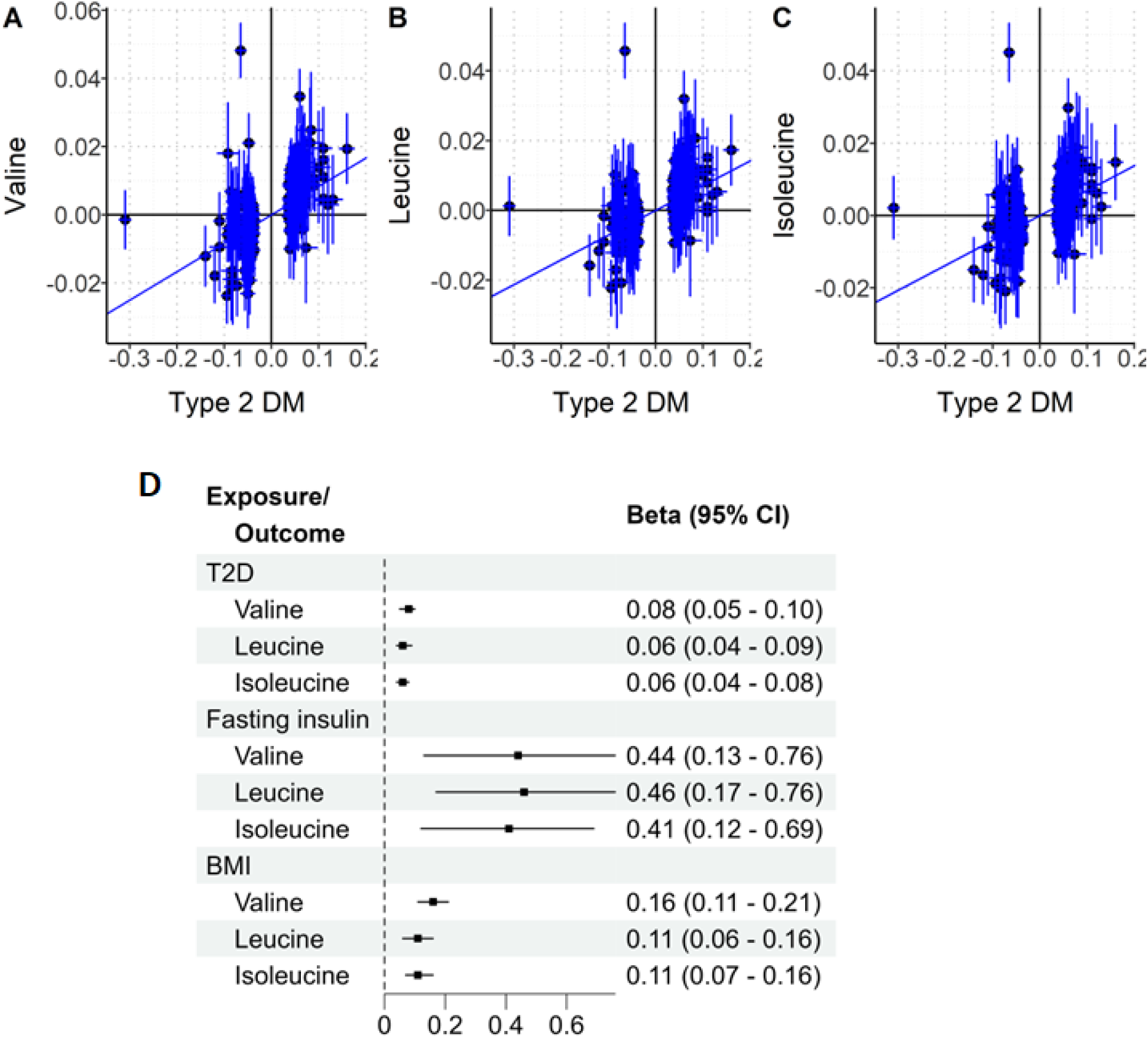
Associations between SNP instruments for metabolic phenotypes and BCAAs. (A-C) Scatter plots showing of the associations between genetic instruments for type 2 diabetes (n=95 SNPs) and valine, leucine and isoleucine. The lines represent the association based on the IVW method. (D) Forest plot summarizing the associations by the inverse variance weighted (IVW) method between genetic instruments for type 2 diabetes, fasting insulin (n=34 SNPs) and body mass index (BMI) (n=69 SNPs) and valine, leucine and isoleucine.

We next constructed instrumental variables for type 2 diabetes (n=95 SNPs) to determine whether the type 2 diabetes was associated with altered BCAA levels. There were significant positive associations with each BCAA (valine: beta=0.08 per standard deviation change in levels per log-odds change in type 2 diabetes, [95% CI: 0.05 - 0.10], p=1.8×10^−9^), (leucine: beta= 0.06 [0.04 - 0.09], p=4.5×10^−8^) and isoleucine (beta= 0.06 [0.04 - 0.08], p=2.8×10^−8^) (**Figure 5**, **Supplemental Table 6**). Sensitivity analyses showed consistent results. Similar significant positive associations were seen when testing associations with with BCAA levels measured in the KORA study (**Supplementary Figure 2, Supplemental Table 7**). We tested associations with BCAA levels in METSIM. In this set, all participants with prevalent or incident type 2 diabetes were excluded from the BCAA GWAS, which would be expected to attenuate or eliminate an association with type 2 diabetes if the association was due to this disease. As expected, while the directions for all associations were consistent with those observed in the UK Biobank and KORA sets, they were not significant for valine and leucine and nominally significant for isoleucine (p=0.03) (**Supplementary Figure 3, Supplemental Table 8**).

**Figure 5:**
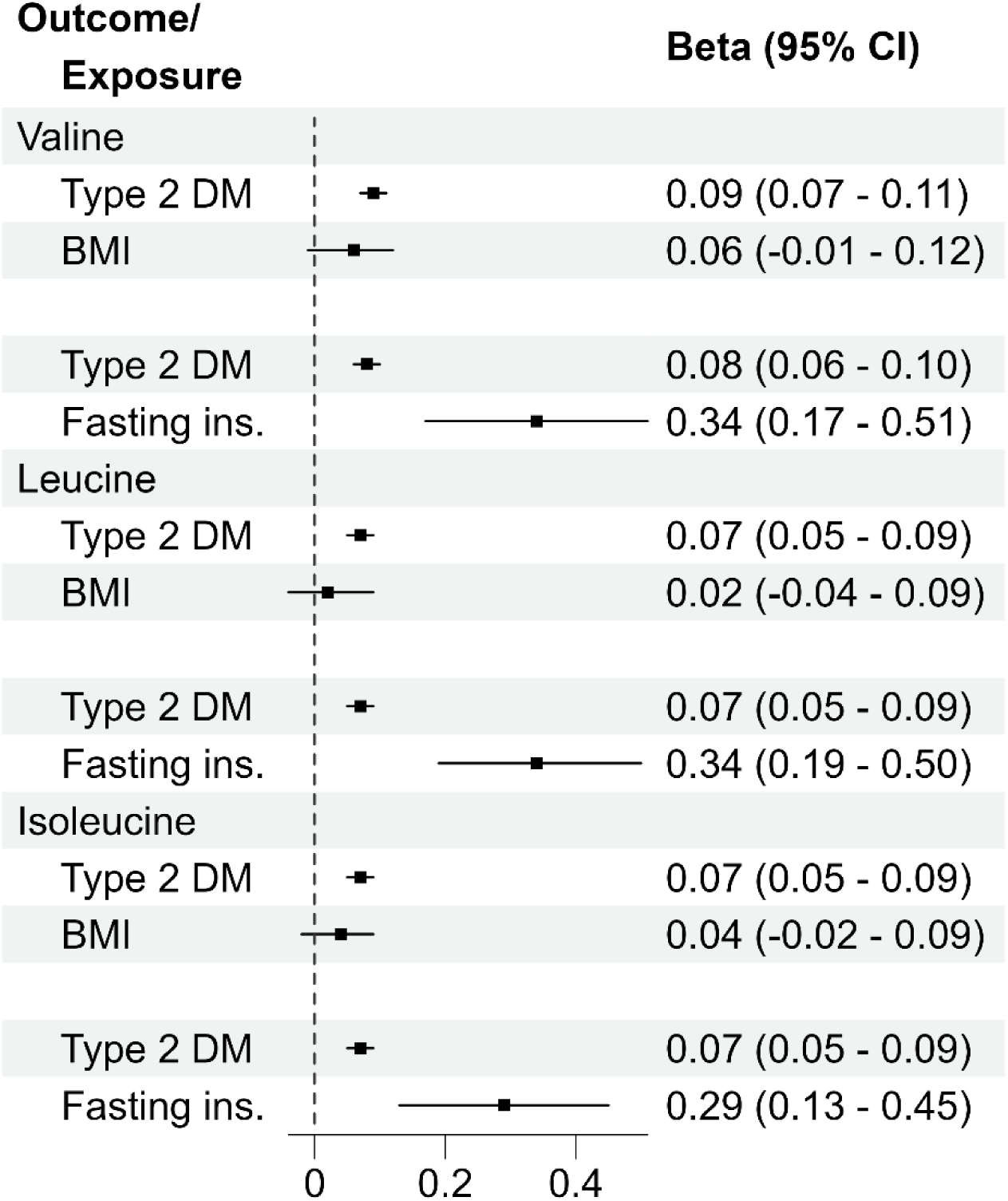
Multivariable MR associations between type 2 diabetes and BCAAs. Forest plots summarizing the multivariable MR associations by the inverse variance weighted (IVW) method between genetic instruments for type 2 diabetes and valine, leucine and isoleucine, adjusted for either BMI or fasting insulin.

The SNPs associated with BCAA levels are associated with insulin regulation and adiposity, suggesting that the BCAAs may be up-stream of these traits. To test this hypothesis, we constructed instrumental variables for fasting insulin levels (n=34 SNPs) and BMI (n=69 SNPs). Similar to type 2 diabetes, there were significant positive associations with each phenotype and each BCAA (**Supplemental Figure 4**, **Supplemental Table 9**). In sensitivity analyses the MR-Egger results for fasting insulin demonstrated discordant results with wide confidence intervals (**Supplemental Table 9**). However, the intercept p-values were not significant. MR-PRESSO analysis did not show significant distortion (p>0.05) after removing outliers. It has previously been observed the MR-Egger can produce unreliable discordant estimates with wide confidence intervals in the presence of SNPs with a direct effects on the outcome.^28,32^ This was the case here, as, for instance, excluding 2 outlying SNPs identified by MR-PRESSO brought the MR-Egger estimate (0.61) in line with the IVW (0.60) and weighted median (0.70) estimates for leucine. In sum, these results suggest that BCAA levels are higher with higher type 2 diabetes risk, fasting insulin levels and BMI.Multivariable MR was used to determine whether the associations with type 2 diabetes was independent of BMI and levels. Type 2 diabetes was associated with each BCAA, independent of BMI and fasting insulin levels (**Figure 5**, **Supplemental Table 8**). However, associations between BMI and the BCAAs were not significant (p>0.05) after adjustment for type 2 diabetes.

## DISCUSSION

We constructed and validated genetic instruments for the BCAAs valine, leucine and isoleucine and tested whether they were associated with type 2 diabetes using standard MR approaches. While the genetic instruments robustly associated with positive control phenotypes, they did not associate with type 2 diabetes. This contrasts with a prior MR study which demonstrated associations for each BCAA.^12^ In line with prior studies, we found that fasting insulin and BMI were positively associated with BCAA levels.^13,33^ In addition, we found that genetic instruments for type 2 diabetes were also robustly associated with BCAA levels, suggesting that the BCAAs are biomarkers, rather than causal mediators of type 2 diabetes. Multivariable MR suggested that the associations with BMI are likely mediated through increasing type 2 diabetes risk.

We did not replicate findings from a prior study that observed associations between genetic instruments of the BCAAs and type 2 diabetes.^12^ There are important differences between that study and this one. The BCAA predictors in that study were heavily weighted with SNPs in high linkage-disequilibrium around the *PPMK1* gene, whose gene product regulates catabolism of each of the BCAAs. Their valine and leucine predictors only used SNPs around that gene locus, and thus were only testing for an association based on a single genetic mechanism. Favoring an association was the fact that there was a suggestive association around the *PPMK1* gene in their type 2 diabetes GWAS and the high representation of SNPs in high LD in this region. We did not observe a type 2 diabetes association at the *PPMK1* locus in the DIAGRAM Consortium GWAS data set. Their isoleucine predictor additionally included SNPs around the *TRMT61A, CBLN1* and *DDX19A*, but not *GCKR*. We did not observe associations and found directional inconsistency between the BCAAs and their lead SNPs around the *TRMT61A* and *CBLN1* genes in the UK Biobank and METSIM cohorts. Thus, we could not replicate their findings due to these non-replicating SNP associations related to isoleucine and type 2 diabetes.

We used a larger set of SNPs measuring diverse genetic mechanisms associated with levels of BCAAs in plasma than prior studies. In general, MR results are most robust when the genetic instruments measure a broad set of independent genetic mechanisms associated with an exposure, as this decreases the likelihood of spurious associations that can result from examining a small number of genetic loci.^26^ Attesting to the validity of the BCAA genetic instruments, we found that the SNP effect sizes corresponded to those measured in an independent cohort. When testing for the type 2 diabetes association, we employed a two-sample design to reduce the chance of identifying associations based on reverse causality and we performed a number of sensitivity analyses to ensure that outlying SNPs were not obscuring an association. Thus, we believe these findings more accurately describe the genetic relationship between the BCAAs and type 2 diabetes.

The SNPs associated with the BCAAs highlight a number of genetic mechanisms influencing circulating BCAA levels, many of which have functions within the mitochondria. In addition to *PPMK1*, the protein product of *BCAT2* transanimates the BCAAs to α-ketoacids while converting α-ketoglutarate (αKG) to glutamate.^34^ The gene products of *GLUD1*, *PRODH* and *GLS2* also impact glutamate metabolism.^35^ *PCCB* contributes to BCAA breakdown by catabolism of propionyl-CoA.^36^ Several genes are related to glucose regulation, including *GCKR* which inhibits glucose metabolism by glucokinase, *SLC2A4 (GLUT4)* which allows glucose entry into cells in response to insulin signaling, and glucose signaling including *MLXIP* which modulates transcription in response to intracellular glucose levels.^37^ *IRS1* and *GRB14* both regulate signaling through the insulin receptor.^38^ There were also genes associated to adiposity including *HNF4G*, a transcription factor found to be associated with BMI in GWAS studies and *RNL3*, which may have a role in appetite regulation.^39–41^ There were also SNPs located at loci that identified genomic regions where the link to BCAAs was not clear including SNPs near *MOK*, *DDX19A-DT, SUOX*^42^ and *DHODH*.

Collectively, the SNP associations support the hypothesis that BCAA levels are influenced not only by gene products directly involved in BCAA catabolism, but also physiological processes related to insulin signaling, glucose regulation and adiposity.^43^ Consistently, we found that genetic instruments related to BMI, fasting insulin levels and type 2 diabetes associated with each BCAA. The associations were in a positive direction, consistent with well-established epidemiological observations. For leucine and isoleucine, the BMI association was not significant after multivariable adjustment, and remained only nominally associated with valine. These results suggest that the BMI association was most likely related to the influences of adiposity on insulin resistance and penetrance of type 2 diabetes.^44^

There are limitations to the current study. False positive associations can arise from the use of weak genetic instruments or the use of pleiotropic SNPs that associate with a risk factor through mechanisms unrelated to the exposure. To mitigate this possibility, we used a range of sensitivity analyses to ensure that associations were consistent across a range of assumptions. These findings also conflict with some epidemiological and experimental data, which could suggest that non-genetically determined BCAA levels may contribute to type 2 diabetes and insulin resistance through mechanisms not captured by these studies. The analyses also relied on existing data that predominantly represents individuals of European ancestries, which limits generalization to other ancestry groups.

In summary, we did not find evidence supporting a causal role for BCAAs on type 2 diabetes risk. Rather, these data support the hypothesis that the BCAAs levels are increased as a consequence of diabetes and insulin resistance.

## AUTHOR CONTRIBUTIONS

JDM and MS had full access to all data and takes responsibility for the integrity of the data and analysis. JDM and JFF provided substantial contributions to the design of the study. MS performed the primary analysis of the BioVU data. MS and DA made substantial contributions to the acquisition of data. JDM, MS and JFF wrote the first draft of the manuscript. All coauthors critically reviewed the manuscript for important intellectual content, provided final approval of the version to be published, and agree to be accountable for some or all aspects of the work presented.

## Data Availability

All data produced in the present work are contained in the manuscript.

---

**Supplementary Figure 1.**
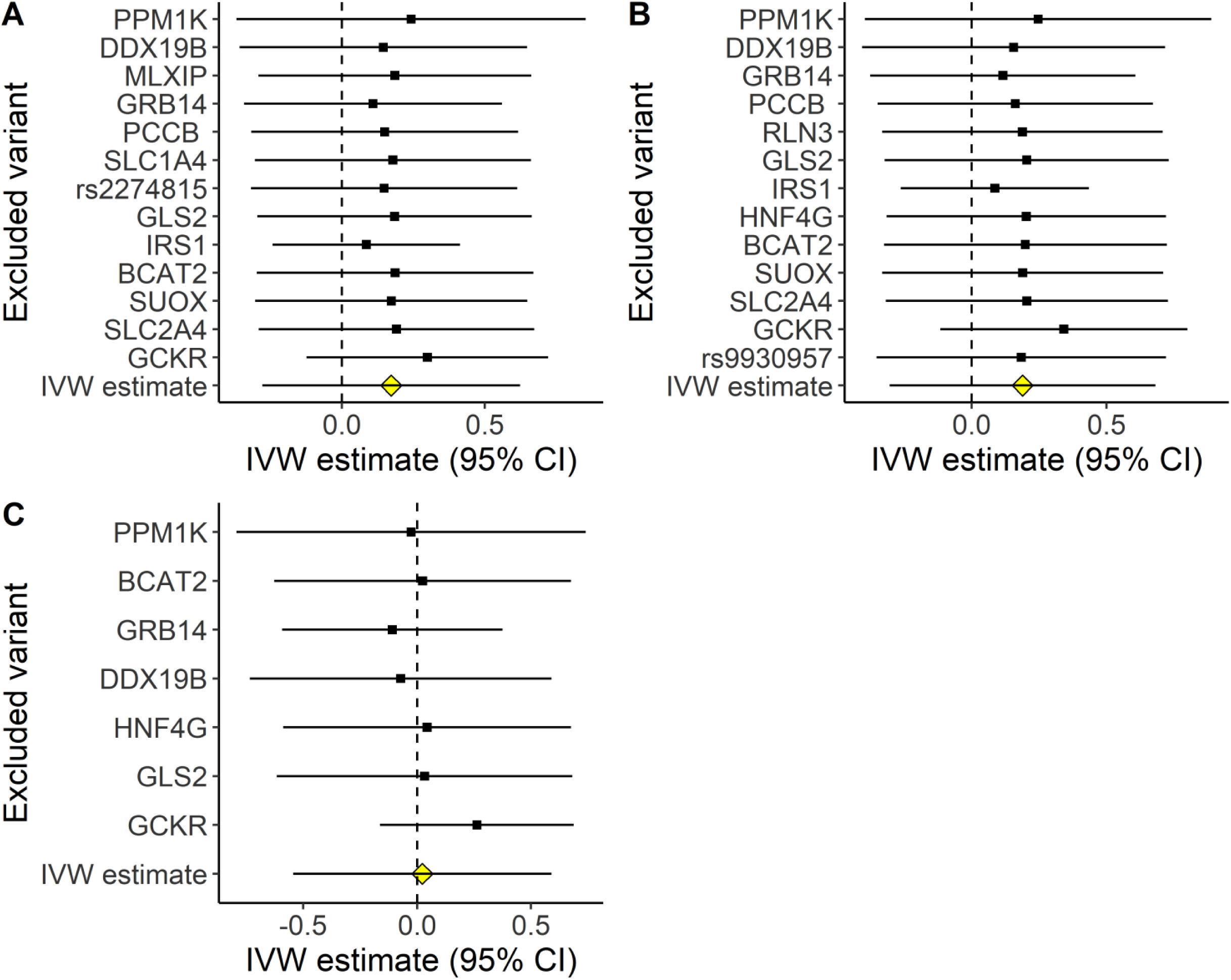
Summary of a leave-one-out analysis for the associations between genetic instruments for (A) valine, (B) leucine and (C) isoleucine and type 2 diabetes. Each figures shows the IVW estimate (95% CI) after exclusion of the indicated SNP. Note that the name of the closest gene was substituted for the SNP name, when available.

**Supplementary Figure 2.**
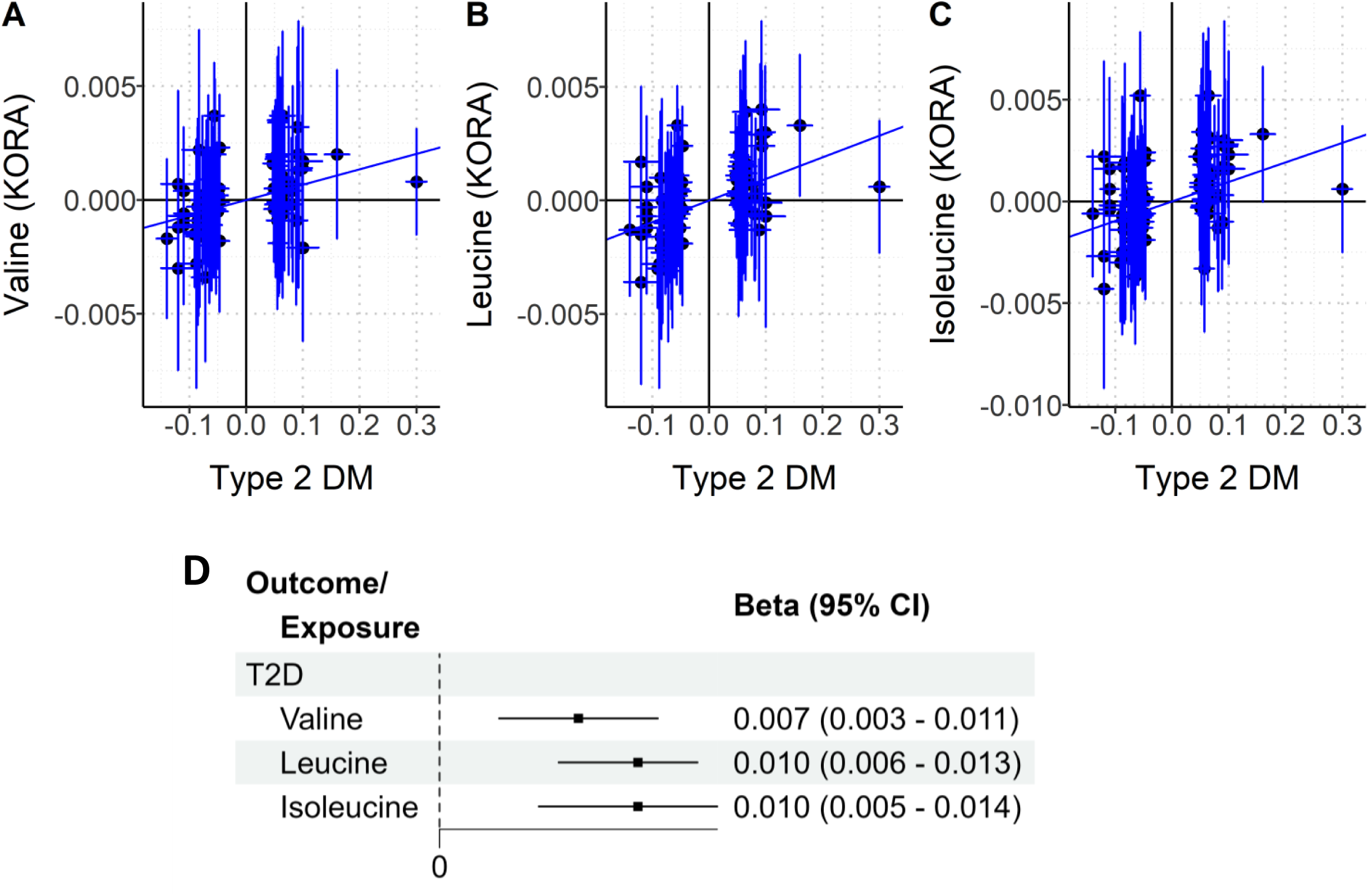
Associations between SNP instruments for type 2 diabetes and BCAAs measured in the KORA population. (A-C) Scatter plots showing of the associations between genetic instruments for type 2 diabetes (n=90 SNPs) and valine, leucine and isoleucine. The lines represent the association based on the IVW method. (D) Forest plot summarizing the associations by the inverse variance weighted (IVW) method between genetic instruments for type 2 diabetes and valine, leucine and isoleucine.

**Supplementary Figure 3.**
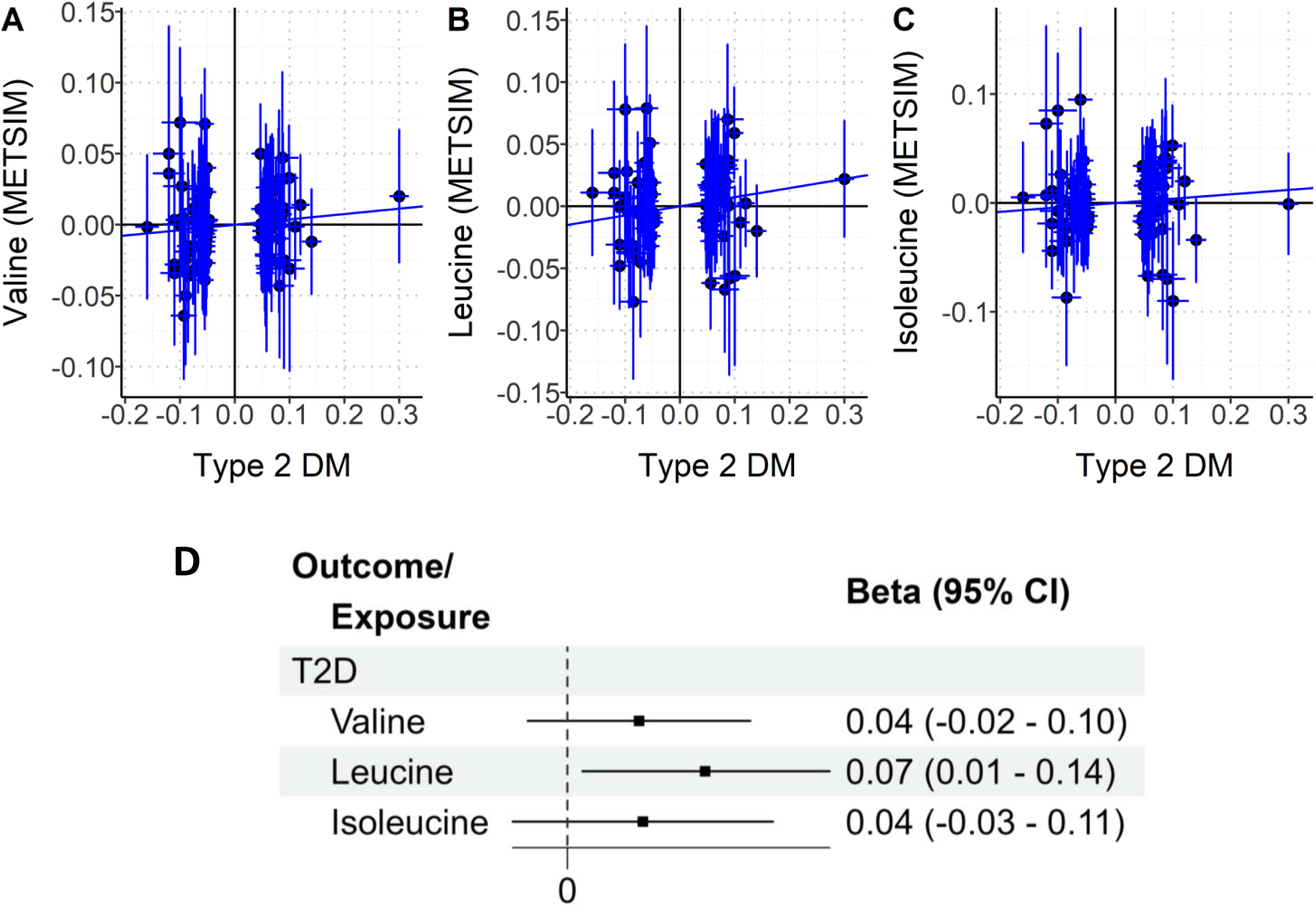
Associations between SNP instruments for type 2 diabetes and BCAAs measured in the METSIM population. (A-C) Scatter plots showing of the associations between genetic instruments for type 2 diabetes (n=95 SNPs) and valine, leucine and isoleucine. The lines represent the association based on the IVW method. (D) Forest plot summarizing the associations by the inverse variance weighted (IVW) method between genetic instruments for type 2 diabetes and valine, leucine and isoleucine.

**Supplementary Figure 4.**
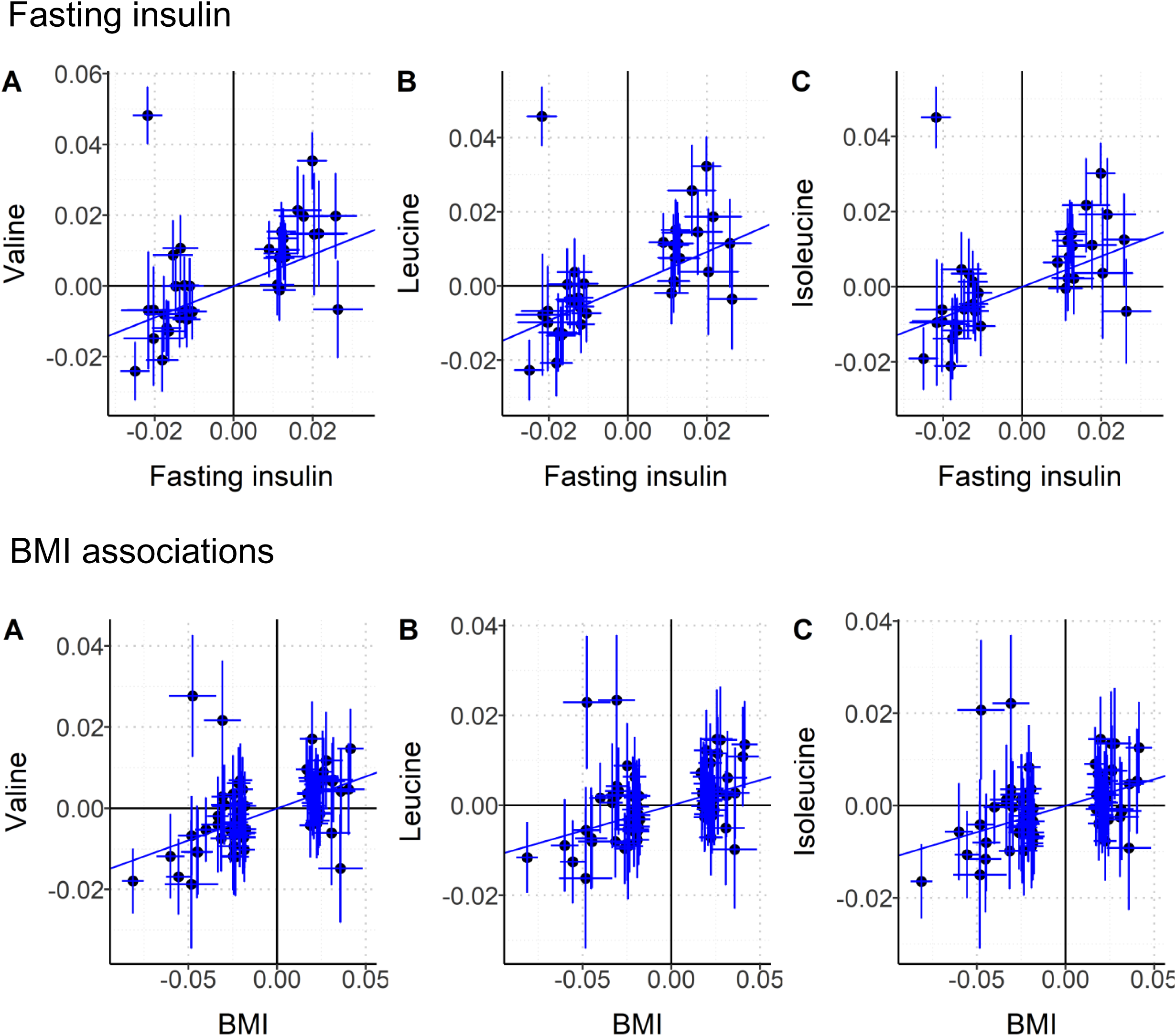
Associations between SNP instruments for fasting insulin or BMI, and the BCAAs measured in the UK biobank population. Scatter plots showing of the associations between genetic instruments for fasting insulin (top panels) and BMI (bottom panels) and each BCAA. The lines represent the association based on the IVW method

